# Bronchoscopy on Intubated COVID-19 Patients is Associated with Low Infectious Risk to Operators

**DOI:** 10.1101/2020.08.30.20177543

**Authors:** Catherine A. Gao, Joseph I. Bailey, James M. Walter, John M. Coleman, Elizabeth S. Malsin, A. Christine Argento, Michelle H. Prickett, NU COVID Investigators, Richard G. Wunderink, Sean B. Smith

## Abstract

**Background:** The coronavirus disease 2019 (COVID-19) pandemic raised concern for exposure to healthcare providers through aerosol generating procedures, such as bronchoalveolar lavage (BAL). Current society guidelines recommended limiting use of BAL to reduce operators’ risk for infection, yet data on the infection rate for providers after BAL is sparse. Since March 2020, our institution used a modified protocol to perform over 450 BALs on intubated COVID-19 patients. We therefore sought to describe the infectious risks to providers associated with BAL.

**Methods:** Fifty-two pulmonary and critical care faculty and fellows at our tertiary-care, urban medical center were surveyed. Survey participants were asked to provide the number of BALs on COVID-19 patients they performed, the number of weeks they worked on intensive care unit (ICU) services with COVID-19 patients, and the results of any SARS-CoV-2 testing that they received. Participants were asked to assess the difficulty of BAL on intubated COVID-19 patients as compared to routine ICU BAL using a numeric perceived difficulty score ranging from 1 (easier) to 10 (harder).

**Results:** We received 47 responses from 52 surveyed (90% response rate), with 2 declining to complete the survey. Many respondents (19/45, 42%) spent >5 weeks on an ICU service with COVID-19 patients.

The number of COVID-19 BALs performed by providers ranged from 0 to >60. Sixteen of the 35 providers (46%) who performed COVID-19 BALs underwent at least one nasopharyngeal (NP) swab to test for SARS-CoV-2, but none were positive. Twenty-seven of the 35 providers (77%) who performed COVID-19 BALs underwent SARS-CoV-2 serology testing, and only one (3.7%) was positive. Respondents indicated occasionally not being able to follow aerosol-minimizing steps but overall felt BALs in COVID-19 patients was only slightly more difficult than routine ICU BAL.

**Discussion:** At a high-volume center having performed >450 BALs on intubated COVID-19 patients with aerosol-limiting precautions, our survey of bronchoscopists found no positive NP SARS-CoV-2 tests and only one positive antibody test result. While the optimal role for COVID-19 BAL remains to be determined, these data suggest that BAL can be safely performed in intubated COVID-19 patients if experienced providers take precautions to limit aerosol generation and wear personal protective equipment.

## INTRODUCTION

The coronavirus disease 2019 (COVID-19) pandemic has caused tremendous amounts of morbidity and mortality. Bacterial co-infections in viral pneumonia have been well described, especially in the setting of influenza pandemics, and have been associated with significant mortality (1). Preliminary studies from COVID-19 have also shown found bacterial co-infection, with especially high rates in those with fatal outcomes (2), suggesting that identification and appropriate treatment of ba(2)cterial co-infection may improve outcomes (3). Preliminary data on a subset of our population found high co-infection rates (4). Given the long average duration of ventilation in COVID-19 patients (5), they are also at high risk for secondary infection with nosocomial bacterial organisms (6).

Therefore, an accurate diagnosis of bacterial superinfection of ventilated COVID-19 patients is important for management, especially to guide appropriate antibiotic stewardship (7, 8). Bronchoalveolar lavage (BAL) with quantitative cultures can be a helpful diagnostic test in the management of severe pneumonia (9). We have a longstanding practice of examining lower respiratory tract samples with rapid diagnostic platforms to inform the diagnosis and treatment of severe pneumonia, as well as research the alveolar microenvironment in severe pneumonia (4, 10).

The standard in our intensive care unit (ICU) had been nonbronchoscopic BAL performed by respiratory therapists, with occasional bronchoscopic BAL for more challenging cases. Since the primary method of severe acute respiratory syndrome coronavirus 2 (SARS-CoV-2) transmission is aerosolization of respiratory droplets (11), nonbronchoscopic BAL was felt to be contraindicated. Endotracheal aspirates have been shown to be inferior to BAL (12, 13). When done correctly for diagnostic purposes, this maneuver also involves a long break in the closed ventilator circuit (14, 15), and thus is not preferred. Bronchoscopic BAL was therefore felt to be the safest procedure for determining the presence and etiology of bacterial infections in intubated COVID-19 patients.

There is concern that bronchoscopy exposes healthcare workers by generating aerosols (16). Society and panel guidelines discourage routine use of bronchoscopy in patients with COVID-19, though this is only based on expert opinion (17, 18). A paucity of data informs the infectious risk to providers performing BAL in COVID-19 patients. In a single-center report of 101 COVID-19 BALs, one of two bronchoscopists became sick with COVID-19 and had to be replaced by a third, who remained uninfected for the duration of the study (19). Some studies where COVID-19 BAL was performed mentioned low provider infection rates, though details are not provided (20).

We therefore designed a safety protocol for BALs in patients intubated with respiratory failure from known or suspected COVID-19. To date in our tertiary-care, urban academic medical ICU, we have performed over 450 bronchoscopies on such patients. Given our high volume of COVID-19 BALs, we designed the following study to assess the incidence of SARS-CoV-2 infection and seropositivity among bronchoscopists.

## METHODS

We surveyed all clinical faculty and fellows in the Pulmonary and Critical Care Division at Northwestern Memorial Hospital (NMH) in Chicago, IL (Survey available as Supplementary 1.) Survey participants were asked to provide the number of COVID-19 BALs that they personally performed or directly participated in, the number of weeks they cared for ICU patients with COVID-19, and the results of any SARS-CoV-2 testing that they had received. Participants were asked to assess the difficulty of BALs on intubated COVID-19 patients compared to routine ICU BALs using a numeric perceived difficulty score ranging from 1 (easier) to 10 (harder). They were also queried regarding COVID-19 exposures outside of work. No identifiers were collected, and all respondents were offered the choice to decline to participate. We received permission from the Vice Dean of Education and our Program Director to survey trainees. This study was reviewed and deemed exempt by our institution’s Institutional Review Board.

### SARS-CoV-2 testing

Nasopharyngeal (NP) testing for SARS-CoV-2 by polymerase chain reaction (PCR) (Becton and Dickinson, Becton, Dickinson and Company, Franklin Lakes, NJ; Cepheid, Sunnyvale, CA; and an institutionally-developed platform) was offered to providers *ad hoc* at the discretion of our hospital’s infection control team, either in response to providers’ symptoms, after a known exposure without PPE, or routine pre-procedure screening for the provider’s own medical care. Serology (Abbott Architect SARS-CoV IgG, Abbott Park, IL) testing was offered to all medical staff by NMH starting in June 2020.

### COVID-19 bronchoscopy protocol (full protocol available in Supplementary 2)

Bronchoscopies were performed in the ICU on intubated patients with respiratory failure from either known or suspected COVID-19. The decision to perform bronchoscopy was at the discretion of the ICU team, in some cases to establish a diagnosis or in others to exclude bacterial co-infection. Bronchoscopies were performed by a pulmonary ICU attending, interventional pulmonology (IP) attending, and/or Pulmonary/IP fellows. Nurses and respiratory therapists were not in the rooms during the actual bronchoscopy. PPE included an N95 mask, eye protection, gloves, gown, and hair protection. Bronchoscopy was performed with a disposable Ambu® aScope™ (Ambu Inc., Columbia, MD) bronchoscope. Patients were sedated per ICU protocols or at the proceduralist’s discretion. Administration of cisatracurium (0.1-0.2 mg/kg) in order to minimize coughing during bronchoscopy was recommended. The endotracheal tube was clamped while the ventilatory circuit was manipulated to accommodate scope placement, and the inspiratory limb of the ventilator was transiently disconnected during the brief break in circuit.

### Statistical Analyses

Survey results were exported from Qualtrics (Qualtrics XM, Provo, UT) and compiled in Microsoft Excel (Version 15.39 for Mac, Microsoft, Redmond, WA). Graphs were generated in GraphPad Prism 8 (version 8.4.3, GraphPad Software LLC), Adobe Illustrator (Version CC for Mac, Adobe Inc, San Jose, CA), and Python using Seaborn v0.10.1. Not all continuous data were normally distributed, and so median values with interquartile ranges (IQR) were reported. Non-parametric analyses included Mann-Whitney-Wilcoxon and Spearman’s rank correlation for continuous variables. Kruskal-Wallis rank testing was used to compare values across multiple categories.

## RESULTS

We surveyed 52 clinical Pulmonary and Critical Care faculty and fellows. Forty-five respondents (90% response rate) agreed to participate: 18 fellows and 27 attendings (Table 1, Figure 1). The majority (35/45, 78%) had performed at least one bronchoscopy. The most common range of bronchoscopies performed by a respondent was 10-30 bronchoscopies, which 14/45 (31%) respondents selected. Two respondents reported performing bronchoscopy without full PPE under emergent situations, and 12 reported performing bronchoscopies without being able to follow the full protocol.

**Table 1:** Survey Results. Pulmonary and critical care fellows and attendings were surveyed as to their weeks on service, number of COVID-19 BALs, and perceived difficulty.

|  | Total | Attendings | Fellows |
| --- | --- | --- | --- |
| Total Surveyed Providers | 52 | 31 | 21 |
| Responded to survey | 47 |  |  |
| Agreed to complete survey | 45 | 27 | 18 |
| Weeks of COVID-19 ICU Service |  |  |  |
| 0 | 9 | 7 | 2 |
| 1 | 2 | 2 | 0 |
| 2 | 3 | 1 | 2 |
| 3 | 4 | 4 | 0 |
| 4 | 8 | 6 | 2 |
| >5 | 19 | 7 | 12 |
| COVID-19 BALs |  |  |  |
| 0 | 10 | 8 | 2 |
| 1 - 10 | 9 | 6 | 3 |
| 10 - 30 | 15 | 9 | 6 |
| 30 - 60 | 5 | 0 | 5 |
| >60 | 6 | 4 | 2 |
| Perceived Difficulty Scores<br>(median, IQR) | 6 (6-7) | 6 (6-7) | 6.5 (5.25-7) |

**Figure 1:**
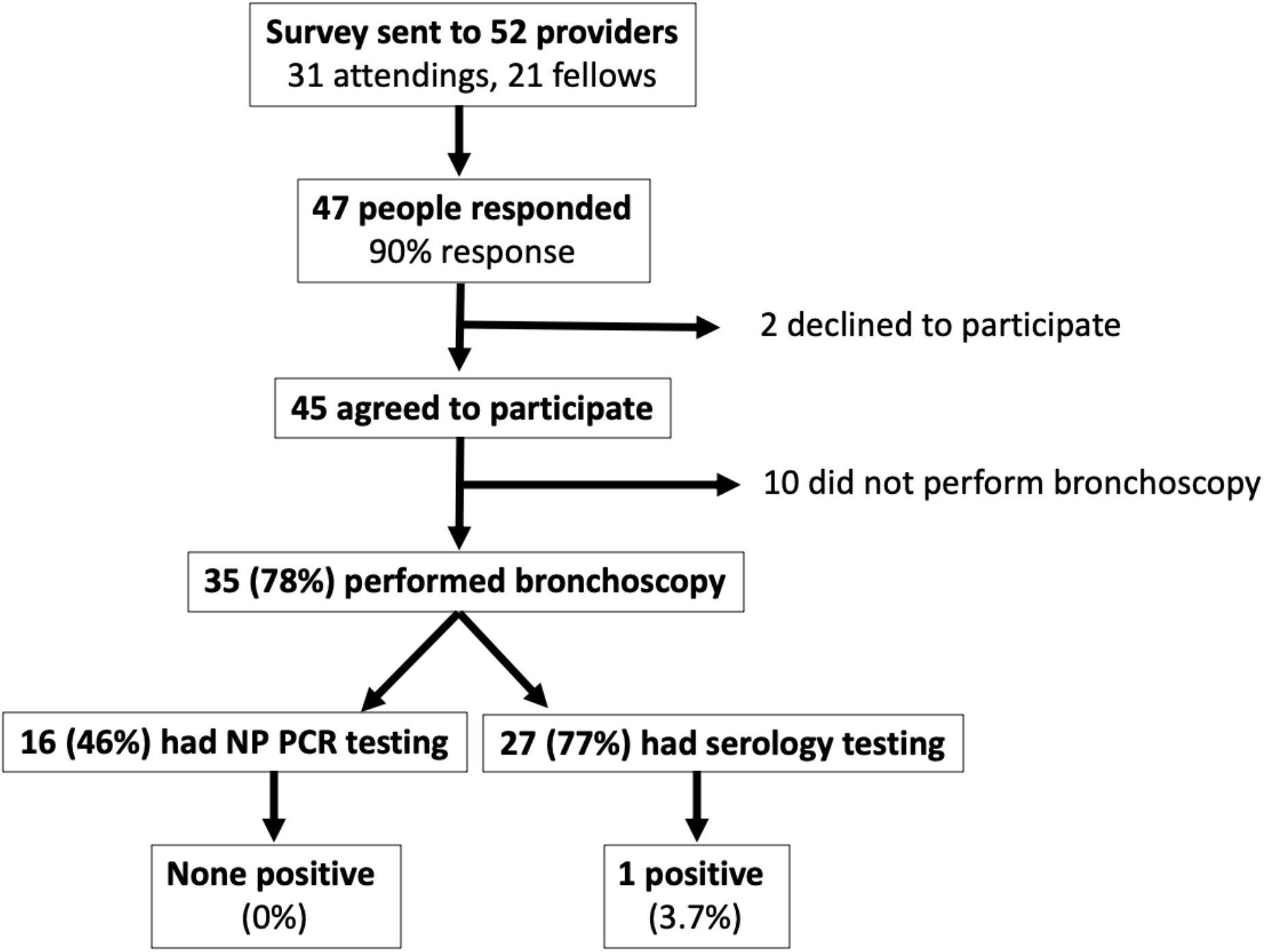
Survey Response Rates and COVID-19 Test Results. Respondents as broken down by those who completed the survey, performed COVID-19 BAL, and their test results.

**Figure 2:**
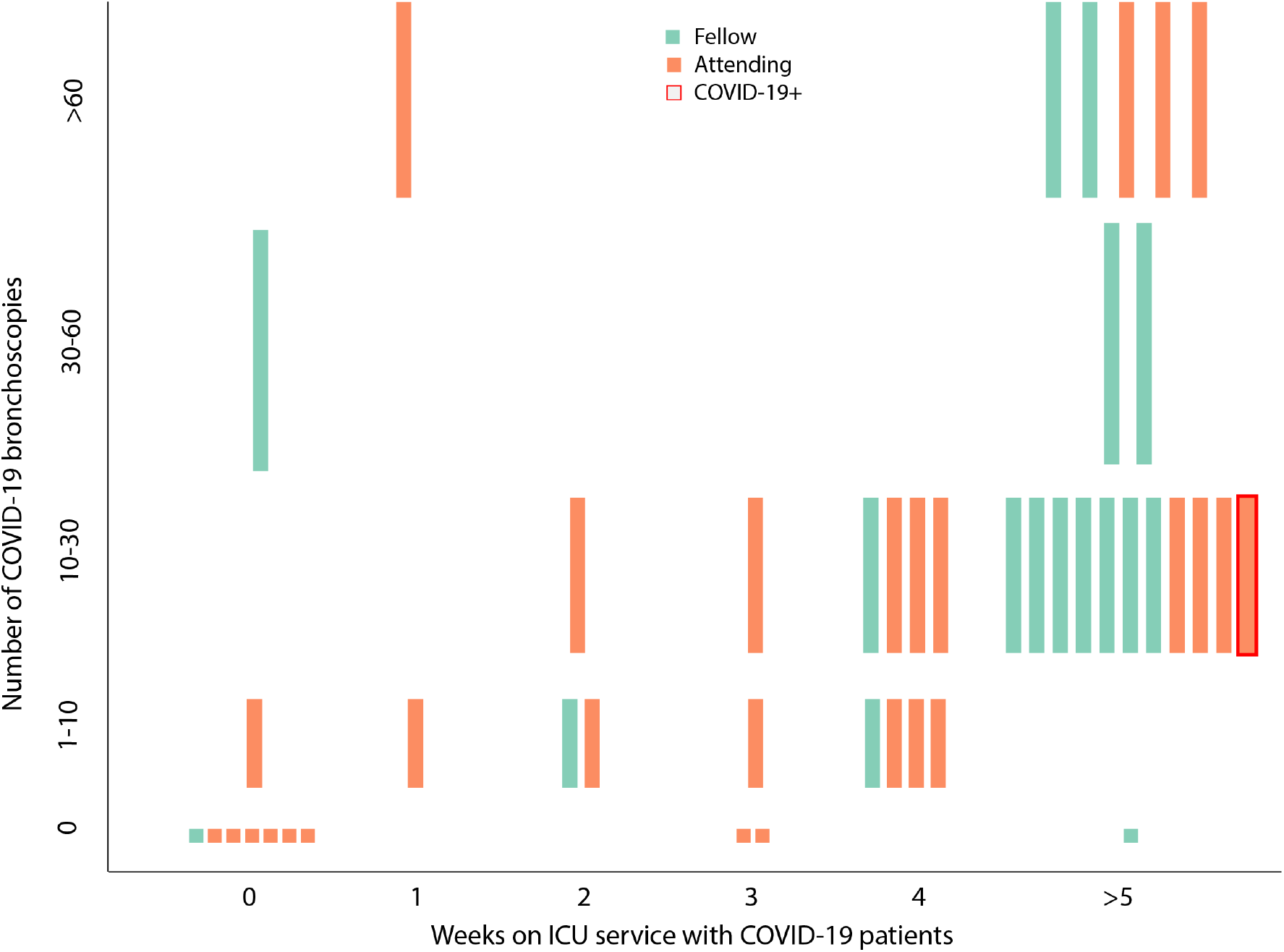
Weeks of COVID-19 ICU Service and Bronchoscopy Volume. The amount of time providers spent on service, and the number of COVID-19 BALs they performed.

Many respondents (42%) spent >5 weeks on an ICU service with COVID-19 patients. The number of weeks on ICU service correlated to bronchoscopy volume (Spearman r=0.66, p<0.05). The overall median perceived difficulty score was 6 (IQR 6-7), with fellows (6.5, 5.75-7 IQR) and attendings (6, IQR 6-7) not differing significantly (p=0.66) (Figure 3). The perceived difficulty score did not correlate with either bronchoscopy volume (p=0.5) or time spent on ICU service with COVID-19 patients (p=0.18). Comments included that respondents felt these bronchoscopies were quite safe given numerous precautions instituted and adequate availability of PPE.

**Figure 3.**
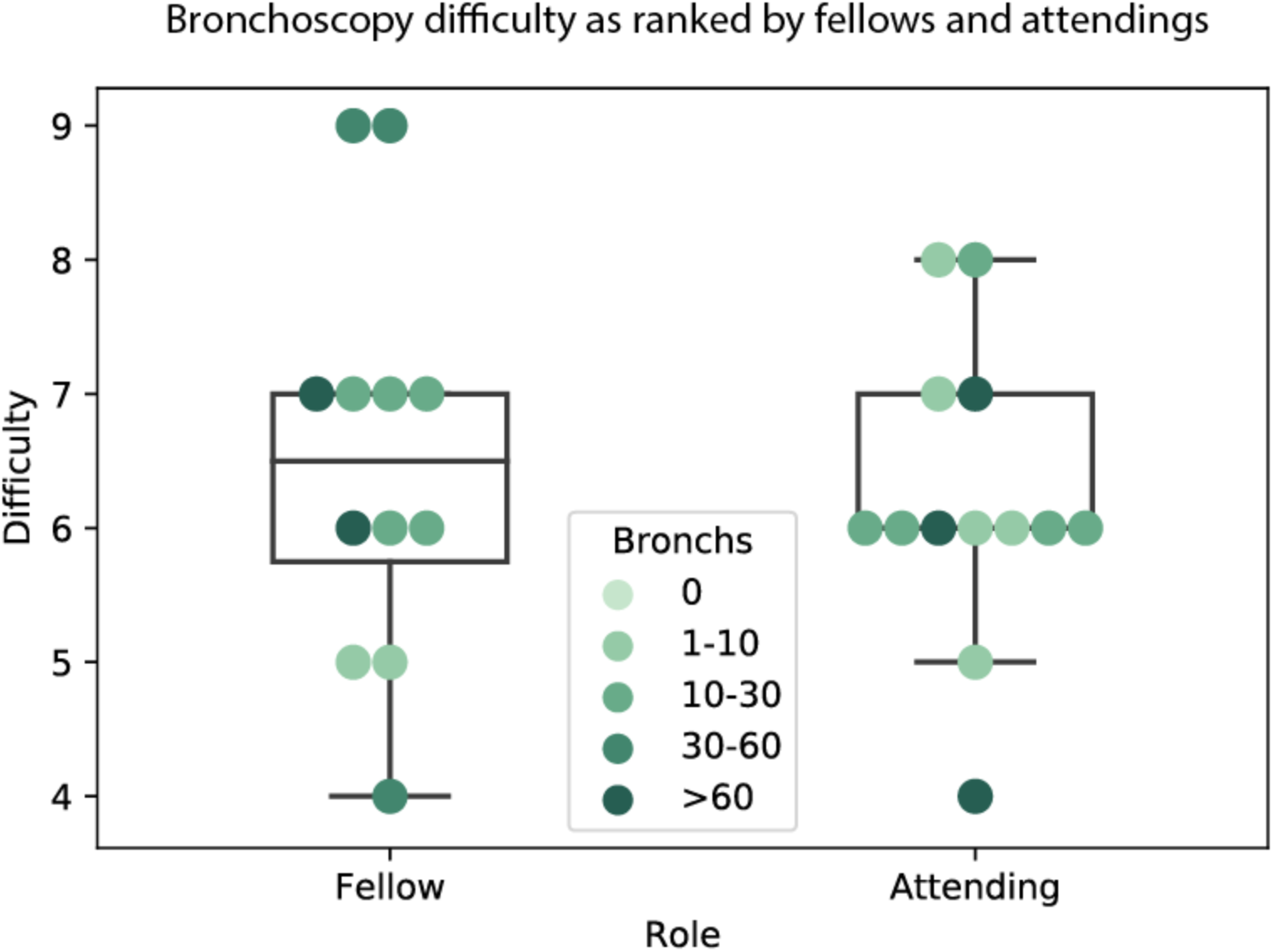
Difficulty as rated by fellows and attendings. Scale of 1 (easier) to 5 (same) to 10 (harder), as compared with non-COVID ICU BAL.

Sixteen of the 35 providers (46%) who performed bronchoscopies underwent at least one nasopharyngeal swab for SARS-CoV-2. No respondents reported a positive test. Five respondents indicated having a respiratory illness that prompted their testing. Eight respondents indicated their SARS-CoV-2 testing was for screening purposes only.

Twenty-seven of the 35 providers (77%) who performed bronchoscopies underwent SARS-CoV-2 serology testing. One individual (3.7%) had two sets of serology testing, with one result being positive and another result being negative. This individual spent >5 weeks on an ICU service with COVID-19 patients, performed 10-30 bronchoscopies, but did not have any febrile respiratory symptoms.

## DISCUSSION

Bronchoalveolar lavage plays an important role in the evaluation of acute respiratory failure in the ICU. Current society guidelines recommend a limited role for bronchoscopy in the care of COVID-19 patients to minimize the potential infectious risk to beside providers of aerosol-generating procedures (17, 18). These guidelines are based on expert opinion and appropriately emphasize provider safety, but data regarding the infectious risk of BAL to providers have been lacking. Our survey is small and limited to a single center, but our data suggest that the risk of transmitting COVID-19 to providers performing BAL may be low. No provider developed COVID-19, and only one had positive serology.

This survey was not a formal study of our specific bronchoscopy protocol. However, we hypothesized that careful technique during bronchoscopy can limit infectious risk. Principles of our protocol included: 1) limiting the number of providers in the room, 2) minimizing aerosol generation by with clamping the endotracheal tube and disconnecting the inspiratory limb of the ventilator during manipulations, 3) minimizing cough by neuromuscular blockade or heavy sedation and instillation of lidocaine into the tracheobronchial tree, and 4) use of a disposable bronchoscope. We also had a small number of highly skilled providers (our IP team) performing a high number of the BALs, ensuring adherence to protocol. Fortunately, most providers felt that the bronchoscopies following the protocol were only slightly more difficult than routine ICU bronchoscopy. We suspect that other centers may have similar protocols, and a more scientific exploration of techniques would be needed to comment on protocol efficacy.

Our group has supported adhering to evidence-based critical care during the COVID-19 pandemic rather than making practice changes solely in response to uncertainties associated with the COVID pandemic (21). Medical provider safety is of paramount importance and precluded relying on our standard of care nonbronchoscopic BAL performed by respiratory therapists. However, our providers already utilize BAL during their routine critical care practice, and were likely to be able to safely continue that practice while using appropriate PPE and minimizing aerosolization. Our results demonstrate that assumption was accurate.

Our study has several limitations. First, this is a retrospective survey of providers and their recollection of practices. We did not directly pull exact bronchoscopy numbers and test results from the electronic medical records so as to respect our colleagues’ privacy, and instead looked for voluntary responses. However, collateral data suggests overestimation of bronchoscopy numbers is very unlikely. Adherence to the modified bronchoscopy protocol was not monitored. We were not able to capture a 100% response rate for the survey but obtained results higher than most physician survey response rates (22).

While serology testing was offered to all providers per hospital protocol, only a subset of providers actually got tested. NP testing was performed *ad hoc* based upon symptoms or exposure. Since neither form of testing was mandated across the survey cohort, both may in fact under-represent the true infectious risk. It is also possible that asymptomatic infections were missed if providers did not seek testing in a short time period after performing said procedures.

In summary, this is the first detailed report of infectious risks from BAL from a large-volume center that routinely incorporates bronchoscopy as part of critical care for patients with COVID-19 respiratory failure. While additional research is needed to inform the optimal use of invasive sampling of the lower respiratory tract to improve outcomes for patients with COVID-19, our data suggest that BAL can be routinely incorporated into the ICU care of these patients with minimal infectious risk to providers.

## Data Availability

Data are available upon request.

## Author contributions

CAG and SBS had full access to all of the data and took responsibility for the integrity of the data and accuracy of data analysis.

Concept and design: CAG, JIB, JMC, AAC, ESM, RGW, SBS

Survey: CAG, JIB, RGW, ACA

Compiling data: CAG, JIB

Statistical analysis: CAG, SBS

Drafting of manuscript: CAG, JIB

Revision of manuscript: CAG, JIB, JMW, JMC, MHP, RGW, SBS Supervision: RGW, ACA, SBS

## Acknowledgements

We thank the many individuals who took care of the COVID-19 patients; this list of physicians and APPs is recognized by their inclusion in the NU COVID Investigators. We thank all the nurses, respiratory therapists, social workers, physical therapists, and providers who cared for these patients.

## Supplementary 1

Survey

Title of Research Study: Provider safety during COVID-19 bronchoscopies

IRB Study Number: STU00213164, version 7-24-20

Principal Investigator: Sean Smith

Supported By: This research is supported by Divison of Pulmonary and Critical Care.

Key Information about this research study: The following is a short summary of this study to help you decide whether to be a part of this study. The purpose of this study is to evaluate the safety of bronchoscopists in COVID-19 patients. You will be asked to complete a survey on the number of bronchoscopies you have performed on COVID patients, the number of COVID tests you have received, and the results of your COVID testing. We expect that you will be in this research study for the duration of this survey.

The primary risk of participation is potentially being identified based on how many bronchoscopies you have performed and the small cohort of participants. The main benefit of participation is publicizing the risks/safety of bronchoscopy on COVID patients. Why am I being asked to take part in this research study?

We are asking you to take part in this research study because you have potentially bronched a COVID-19 patient.

How many people will be in this study? We expect about 40 people here will be in this research study.

What should I know about a research study?

- Whether or not you take part is up to you.
- You can choose not to take part.
- You can agree to take part and later change your mind.
- Your decision will not be held against you.

If you say that “Yes, you want to be in this research,” here is what you will do:You will be asked to complete a survey on the number of bronchoscopies you have performed on COVID patients, the number of COVID tests you have received, and the results of your COVID testing. Will being in this study help me in any way?

We cannot promise any benefits to you from your taking part in this research. However, possible benefits include publicizing the risks/safety of bronchoscopy on COVID patients

Is there any way being in this study could be bad for me?

The primary risk of participation is potentially being identified based on how many bronchoscopies you have performed and the small cohort of participants.

What happens if I do not want to be in this research or if I say “Yes”, but I change my mind later?

Participation in research is voluntary. You can decide to participate or not to participate.

You can decide not to participate in this research or you can start and then decide to leave the research at any time and it will not be held against you. To do so, simply exit the survey. Any data already collected will not be saved.

What happens to the information collected for the research?

Efforts will be made to limit the use and disclosure data gathered to people who have a need to review this information. We cannot promise complete secrecy. Organizations that may inspect and copy your information include the IRB and other representatives of this institution.

This survey is being hosted by Qualtrics and involves a secure connection. Upon receiving results of your survey, any possible identifiers will be deleted. You will be identified only by a unique subject number. All information will be kept on a password protected file only accessible by the research team. The results of the research study may be published, but your name will not be used.

### Data Sharing

De-identified data from this study may be shared with the research community at large to advance science and health. We will remove or code any personal information that could identify you before files are shared with other researchers to ensure that, by current scientific standards and known methods, no one will be able to identify you from the information we share. Despite these measures, we cannot guarantee anonymity of your personal data.

What else do I need to know?

If you agree to take part in this research study, we will provide you with the survey questions.

Who can I talk to?

If you have questions, concerns, or complaints talk to the Principal Investigator Sean Smith, or Catherine Gao

This research has been reviewed and approved by an Institutional Review Board (“IRB”). You may talk to them at (312) 503-9338 or if:

- Your questions, concerns, or complaints are not being answered by the research team.
- You cannot reach the research team.
- You want to talk to someone besides the research team.
- You have questions about your rights as a research participant.
- You want to get information or provide input about this research.

Do you agree to participate?

> I agree or I decline

What is your role?

> Attending or Fellow

How many weeks did you spend on the COVID ICU services?

> 0, 1, 2, 3, 4, >5, other:

How many bronchoscopy procedures did you perform on COVID positive patients? Please provide the exact number if you have this available, or select an estimate from the ranges offered:

> 0, 1-10, 10-30, 30-60, >60, other:

Were you ever tested for COVID via nasopharyngeal swab?

> Was this for symptoms or for screening or in response to symptoms?
>
> If yes, were any of these tests positive?

Were you ever tested for COVID-19 by antibody serology testing?

> If yes, did you have detectable COVID antibodies?

Did you have any febrile respiratory illness during the COVID-19 pandemic (starting Jan 1 2020 to now)? Yes/No

Did you ever perform a bronchoscopy without access to the recommended COVID-19 personal protective equipment – N95, eye protection, gown, and gloves?

If yes, estimate how many procedures were performed without recommended PPE:

Did you ever perform a bronchoscopy on a COVID positive patient without following the droplet minimization steps of clamping the endotracheal tube and disconnecting the ventilator? Yes/No

If yes, estimate how many procedures were performed without following droplet mitigation procedures.

Did you have any other high risk COVID exposures outside of work?

Compared to a typical, non-COVID bronchoscopy in intubated patients, how much more difficult were the COVID bronchoscopies (eg, donning/doffing PPE, clamping ETT, disconnecting inspiratory limb of the ventilator)?

> (1 – much easier, 5 – same, 10 – much more difficult; sliding scale)

Other questions or comments: (free text)

## Supplementary 2

NMH COVID-19 ICU Bronchoscopy protocol

Bronchoscopy in the COVID ICU will be performed for diagnostic and therapeutic purposes, including, but not limited to:

- Diagnostic evaluation of newly intubated patient, including COVID rule-out testing
- Evaluation of possible VAP or superimposed bacterial CAP
- Airway clearance

Bronchoscopy can be performed at the discretion of the ICU attending, but the general policy will be that off-hour bronchoscopy performed by a fellow without an attending should be limited to emergent situations such as mucous plugging. The following protocol will be used for bronchoscopy and will be performed by 1-2 member(s) of the IP service or the ICU attending. The ICU fellow will participate if willing and available (per fellow’s preference).

Given the longer circuit break with NBBAL, these will not be performed on COVID-19 patients.

The nurse will help with printing order labels, pre-procedural sedation including administration of neuromuscular blockade but then will leave the room.

RT may help with gathering equipment but does not need to be present in the room.

Role 1: primary bronchoscopist (fellow, attending, IP attending) – this person is responsible for assessing the clinical situation, consenting the family, ensuring all necessary equipment are ready, ordering lab tests, ensuring adequate sedation, performing the bronchoscopy, cleaning the equipment afterwards, and ensuring samples delivered to the lab.

Role 2: secondary bronchoscopist (supervising IP attending or ICU attending or fellow) – this person will assist the primary bronchoscopist, silence ventilator alarms, assist in circuit manipulation, instill saline for lavage, withdraw BAL, and connect lukens trap.

Order checklist:

- Adequate sedation – goal of RASS -4 if neuromuscular blockade to be used
- Cisatricurium 0.2mg/kg (Pharmacist may often have, otherwise call 9^th^ floor pharmacy)
- Labwork

- BAL cell count and differential
- BAL amylase
- BAL respiratory culture (normal, +/- fungal, AFB per clinician determination)
- Lower Respiratory Tract Panel (BioFire Pneumonia Panel)
- SARS-CoV-2 test (repeat even if status already known, to monitor for clearance/reinfection)
- Cytology, Galactomannan, PJP DFA per clinician determination
- Extra patient label for research specimen

PPE checklist:

- N95 mask (ensure adequate fit) + covering surgical mask
- Gown, gloves (discard after procedure)
- Faceshield or goggles (wipe down after procedure)

Equipment checklist:

- Ambu scope (large/orange if concern for mucus plugging, but check ETT size; regular/green otherwise)
- Ambu tower (ensure adequately charged)
- Drape or chuck to lay down supplies on
- Scope adaptor for ETT
- Clamp for ETT
- Scope lubricant
- Extra suction tubing, if far from the bed
- 4x 30cc syringes (slip tip preferred – if luer lock, ensure slip tip adaptors available; often in the scope bag)
- Normal saline (500cc bottles)
- Lukens trap
- Orange specimen cup + labels
- Research Eppendorf tube + patient sticker
- Extra specimen bags outside room for double-bagging specimens

Procedure steps:

1. Outside the room, Primary Bronchoscopist times out with nursing and ensures consent has been signed, orders placed and labels printed. Sedation adjusted to goal RASS-4 with cisatraciurium 0.2mg/kg administered at provider’s discretion (this is suggested when patient in early/acute phase with likely high viral load and tenuous status but may not be necessary for convalescing patients late in disease stage). FiO2 increased to 100%.
2. As able, Primary Bronchoscopist preps the bronch equipment outside the room

a. Drawing up 30cc normal saline x4
b. Connecting AmbuScope to Monitor
c. Lubricating AmbuScope
d. Pre-loading AmbuScope onto scope adaptor
e. Applying labels to specimen cups and research tube
3. Primary Bronchoscopist and Secondary Bronchoscopist enter room; Nursing and RT available but are outside the room.

a. Primary Bronchoscopist responsible for narrating steps out loud, stands at the side of the patient; Secondary Bronchoscopist stands next to first, closer to the ventilator
b. Primary Bronchoscopist ensures equipment set up, suction connected and functioning, sedation is adequate and vitals are stable to tolerate the procedure
c. Primary Bronchoscopist clamps ETT
d. Secondary Bronchoscopist disconnects inspiratory limb from the vent distal to the filter
e. Primary Bronchoscopist places the adaptor (pre-loaded with the scope) onto the ETT
f. Secondary Bronchoscopist reconnects the inspiratory limb
g. Primary Bronchoscopist unclamps the ETT
h. Primary Bronchoscopist performs inspection, toileting secretions as needed, wedges into target lobe
i. Secondary Bronchoscopist instills saline in 30cc aliquots, 120cc recommended, draws back and discards first 5 cc, draws back more sample if able, then connects the lukens trap (goal >40cc return)
j. Primary Bronchoscopist suctions sample into lukens trap
k. Secondary Bronchoscopist disconnect lukens trap, hooks back up to wall suction
l. Primary Bronchoscopist cleans up any remaining secretions, pulls scope back to edge of adaptor, then clamps ETT
m. Secondary Bronchoscopist disconnects inspiratory limb from the vent distal to the filter
n. Primary Bronchoscopist removes adaptor and scope in one motion
o. Secondary Bronchoscopist reconnects the inspiratory limb
p. Primary Bronchoscopist unclamps the ETT
q. Sample placed into orange specimen cup from lukens trap (have had several break), with 10-15cc placed in Eppendorf tube for research team

i. Samples bagged first in room, then placed in another bag held by someone outside the room
ii. Orange specimen cup + labels delivered to 7^th^ floor lab
iii. Research specimen – The MICU research team can be contacted by phone at 62752 or by pager at 59285. If they are not readily available to pick up the specimen, it can be left in the specimen fridge in the dirty utility room in the 9th floor MICU.
r. Disposable equipment placed in red biohazard bag for disposal
s. Monitor and pole wiped down before leaving room, returned to RT room
t. Primary Bronchoscopist returns FiO2 to pre-procedure level (assuming tolerated) and ensures hemodynamics acceptable
u. Both proceduralists doff PPE and wash hands; surgical mask over N95 should be discarded but N95 can be reused; goggles/faceshields wiped down and reused
v. Primary Bronchoscopist documents procedure note

Special Circumstances:

- Prone positioning: continue with standard procedure
- iNO: continue with standard procedure
- Brushings: sometimes requested by research team, who will provide brushes, brush cutter (wire cutter can also be found in bronch suite), research tube/medium for brush to be cut into

Please do not hesitate to contact the Interventional Pulmonary team with questions.

Protocol updated 8-29-2020 by Catherine Gao, from version 4-5-20 from COVID-ICU-Guidelines.

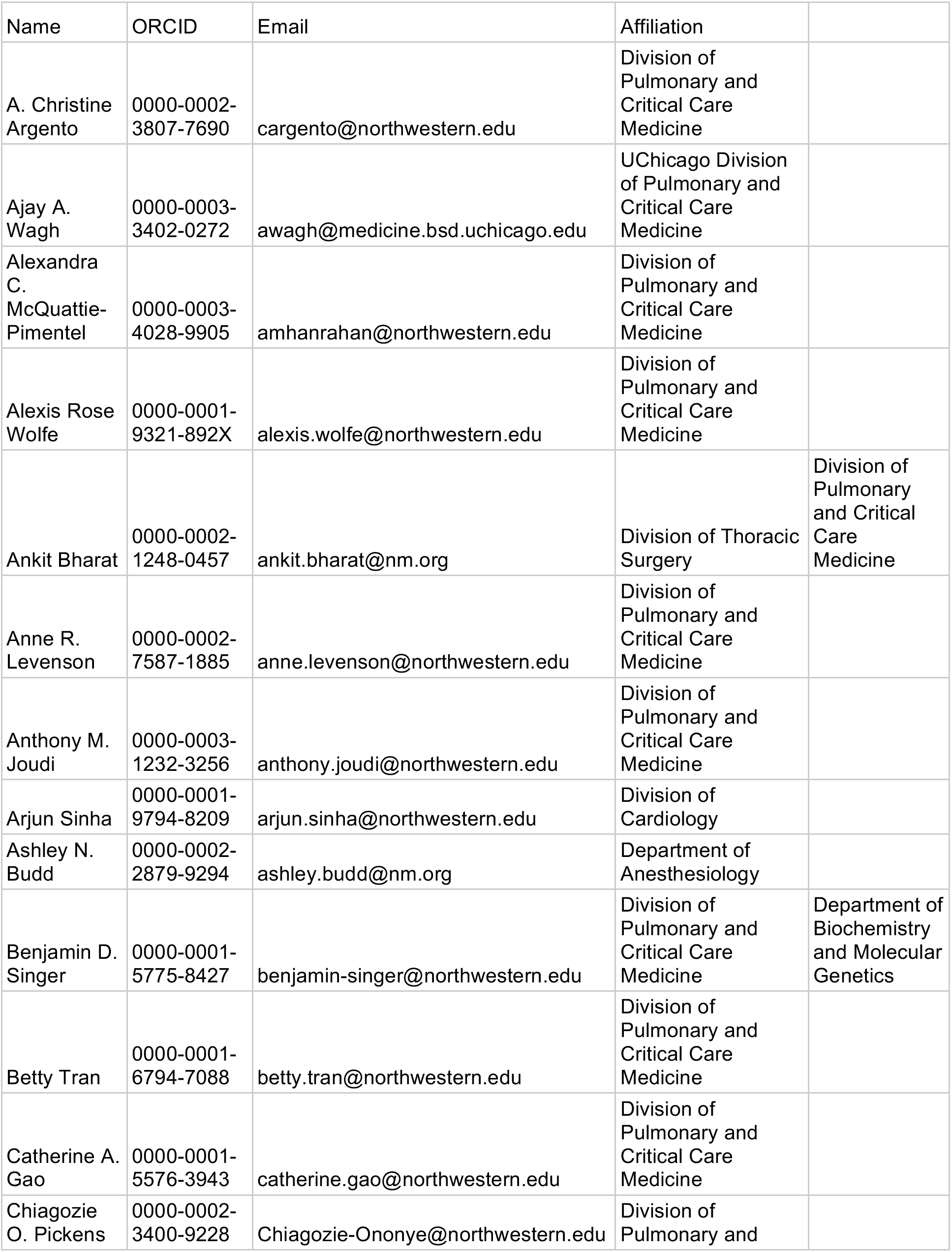
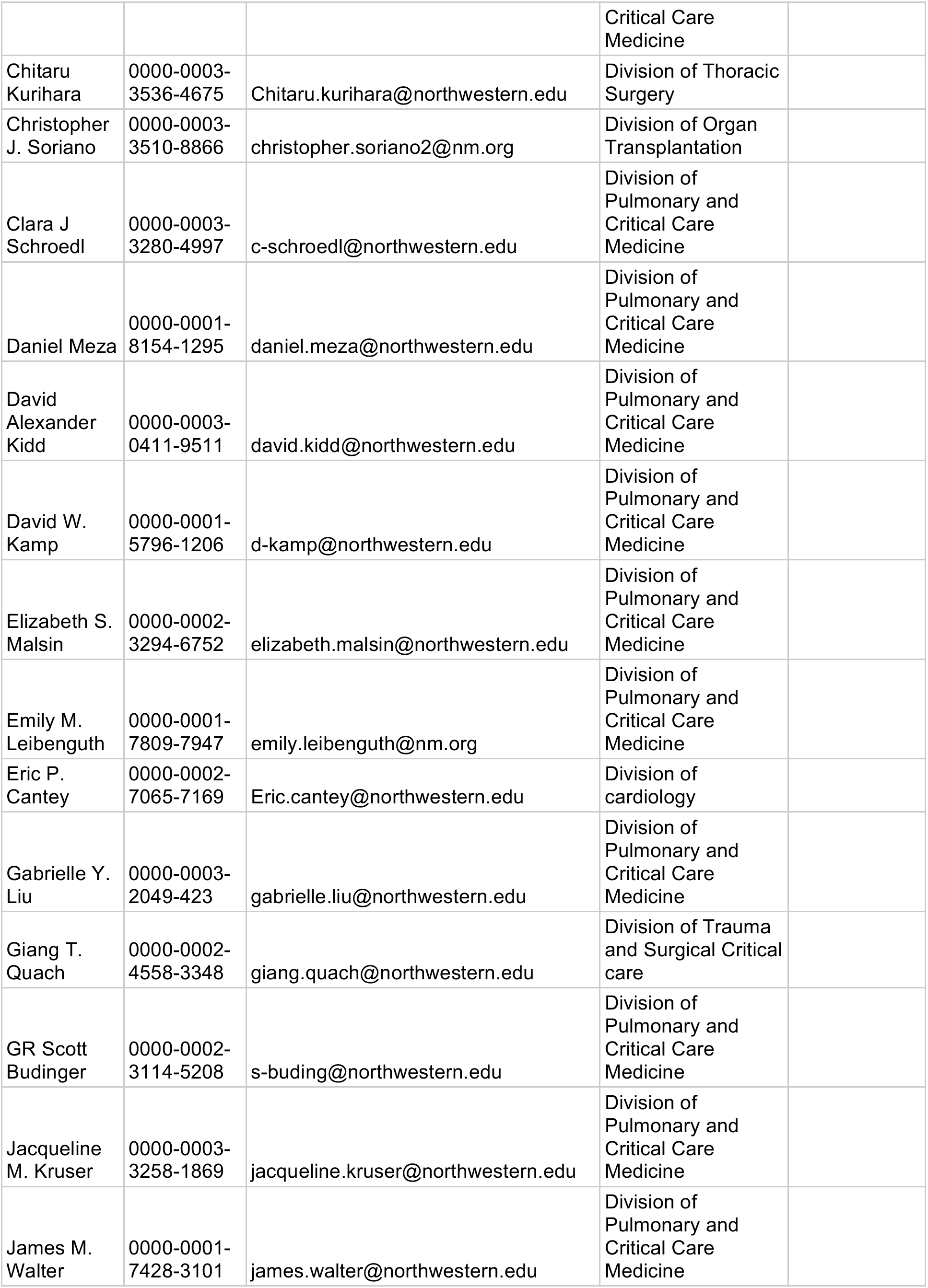
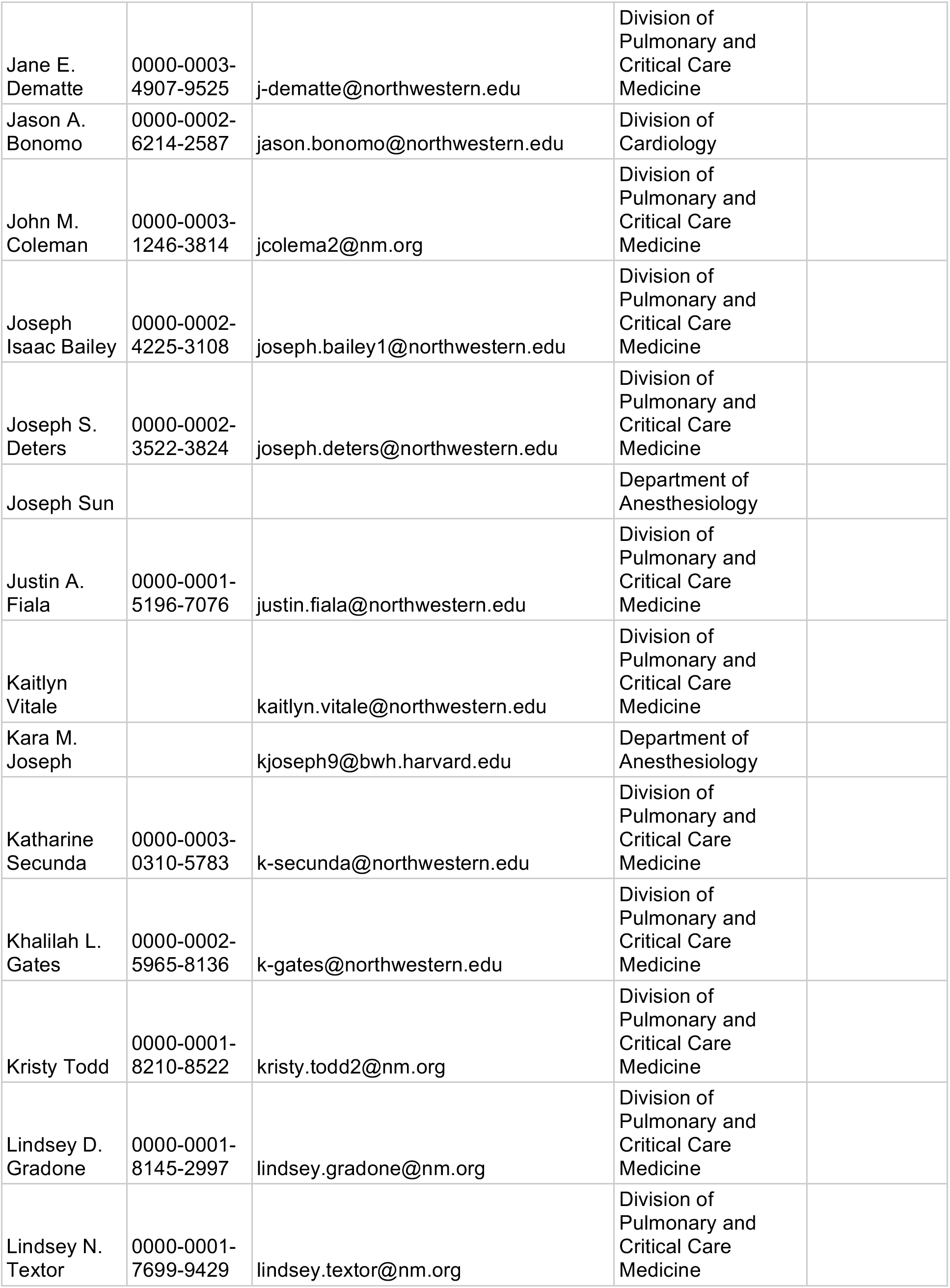
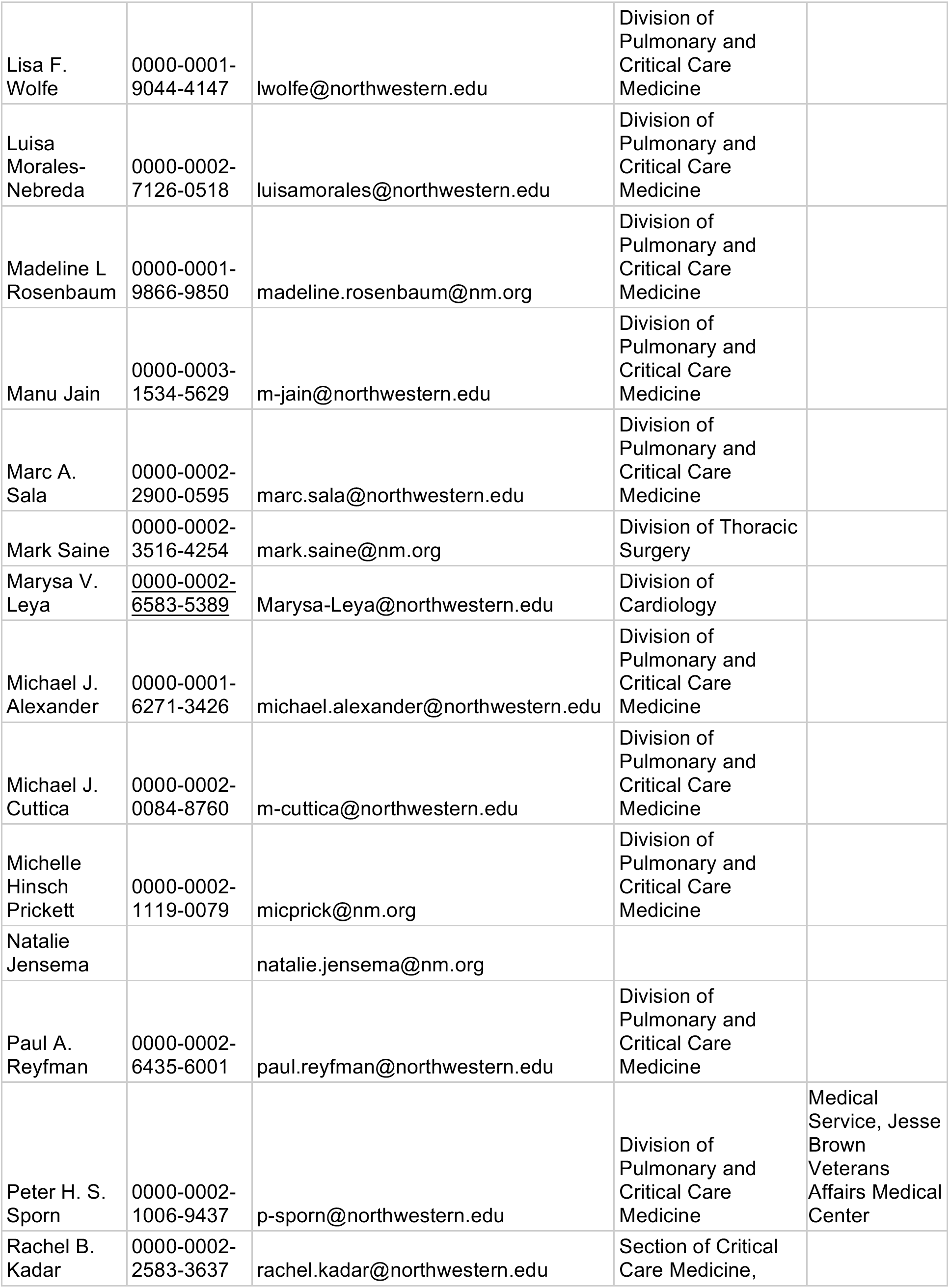
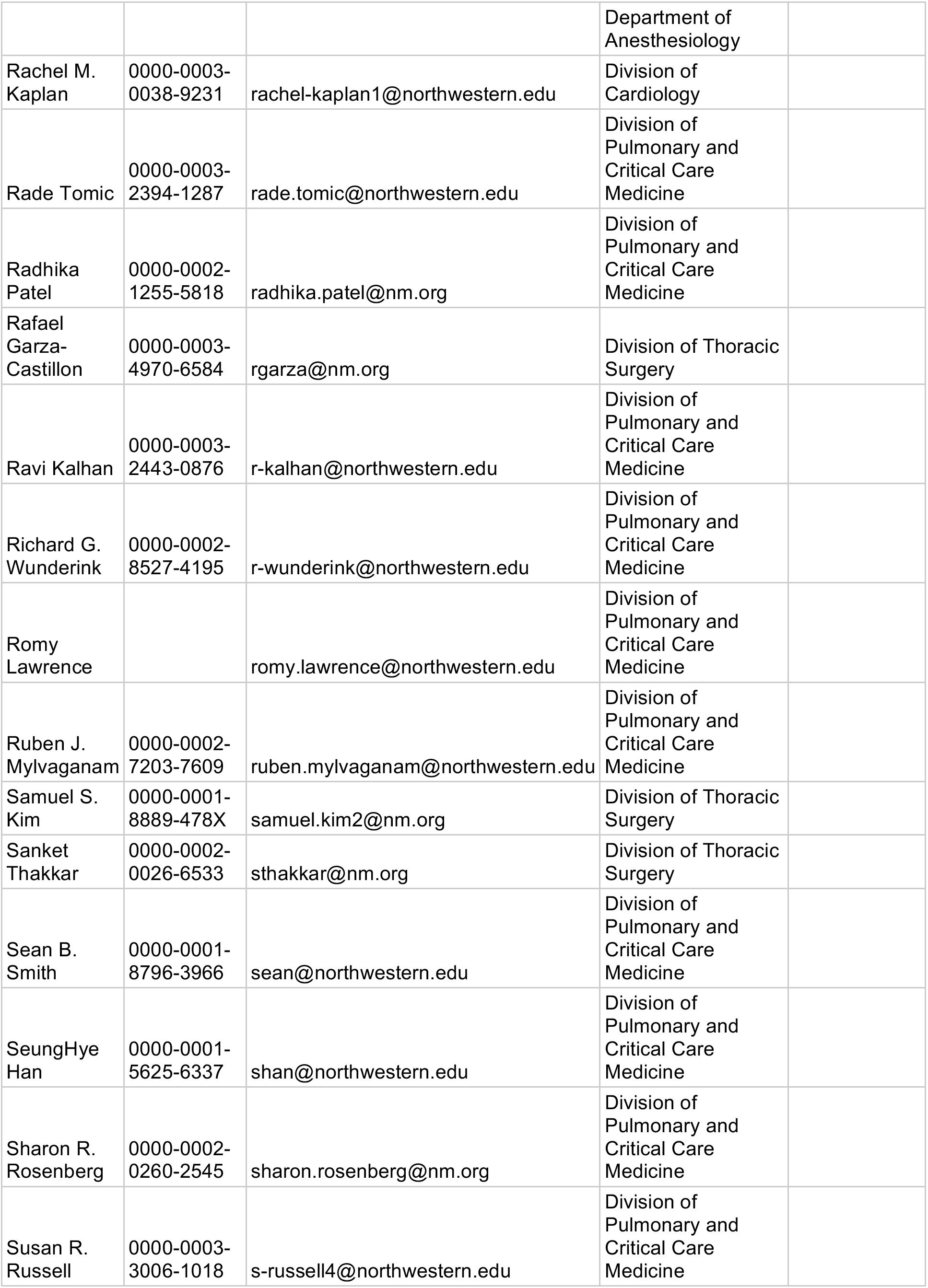
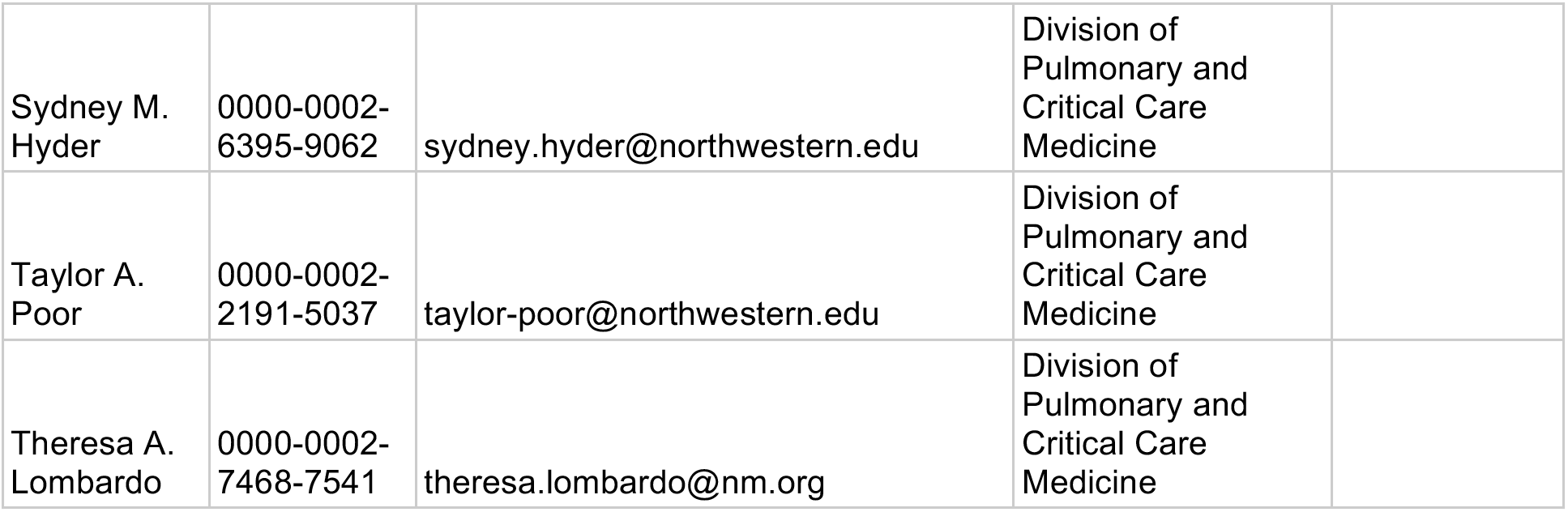
NU COVID Investigators

## Works Cited

1. Martin-Loeches I, Schultz MJ, Vincent J-L, Alvarez-Lerma F, Bos LD, Solé-Violán J, Torres A, Rodriguez A. Increased incidence of co-infection in critically ill patients with influenza. Intensive Care Medicine 2017;

2. Zhou F, Yu T, Du R, Fan G, Liu Y, Liu Z, Xiang J, Wang Y, Song B, Gu X, Guan L, Wei Y, Li H, Wu X, Xu J, Tu S, Zhang Y, Chen H, Cao B. Clinical course and risk factors for mortality of adult inpatients with COVID-19 in Wuhan, China: a retrospective cohort study. The Lancet 2020;

3. Cox MJ, Loman N, Bogaert D, O’Grady J. Co-infections: potentially lethal and unexplored in COVID-19. Lancet Microbe 2020;1:e11.

4. Grant RA, Morales-Nebreda L, Markov NS, Swaminathan S, Guzman ER, Abbott DA, Donnelly HK, Donayre A, Goldberg IA, Klug ZM, Borkowski N, Lu Z, Kihshen H, Politanska Y, Sichizya L, Kang M, Shilatifard A, Qi C, Christine Argento A, Kruser JM, Malsin ES, Pickens CO, Smith S, Walter JM, Pawlowski AE, Schneider D, Nannapaneni P, Abdala-Valencia H, Bharat A, et al. Alveolitis in severe SARS-CoV-2 pneumonia is driven by self-sustaining circuits between infected alveolar macrophages and T cells. 2020;doi:10.1101/2020.08.05.238188.

5. Madhok J, Mihm FG. Rethinking sedation during prolonged mechanical ventilation for COVID-19 respiratory failure. Anesth Analg 2020;doi:10.1213/ANE.0000000000004960.

6. Koenig SM, Truwit JD. Ventilator-associated pneumonia: diagnosis, treatment, and prevention. Clin Microbiol Rev 2006;19:637–657.

7. Pickens CI, Wunderink RG. Principles and Practice of Antibiotic Stewardship in the ICU. Chest 2019;

8. Wunderink RG, Srinivasan A, Barie PS, Chastre J, Cs DC, Douglas IS, Ecklund M, Evans SE, Evans SR, Gerlach AT, Hicks LA, Howell M, Hutchinson ML, Hyzy RC, Kane-Gill SL, Lease ED, Metersky ML, Munro N, Niederman MS, Restrepo MI, Sessler CN, Simpson SQ, Swoboda SM, Guillamet CV, Waterer GW, Weiss CH. Antibiotic Stewardship in the Intensive Care Unit. An Official American Thoracic Society Workshop Report in Collaboration with the AACN, CHEST, CDC, and SCCM. Ann Am Thorac Soc 2020;17.:

9. Torres A, Niederman MS, Chastre J, Ewig S, Fernandez-Vandellos P, Hanberger H, Kollef M, Li Bassi G, Luna CM, Martin-Loeches I, Paiva JA, Read RC, Rigau D, Timsit JF, Welte T, Wunderink R. International ERS/ESICM/ESCMID/ALAT guidelines for the management of hospital-acquired pneumonia and ventilator-associated pneumonia: Guidelines for the management of hospital-acquired pneumonia (HAP)/ventilator-associated pneumonia (VAP) of the European Respiratory Society (ERS), European Society of Intensive Care Medicine (ESICM), European Society of Clinical Microbiology and Infectious Diseases (ESCMID) and Asociación Latinoamericana del Tórax (ALAT). Eur Respir J 2017;50.:

10. Walter JM, Helmin KA, Abdala-Valencia H, Wunderink RG, Singer BD. Multidimensional assessment of alveolar T cells in critically ill patients. JCI Insight 2018;3.:

11. Shereen MA, Khan S, Kazmi A, Bashir N, Siddique R. COVID-19 infection: Origin, transmission, and characteristics of human coronaviruses. J Advert Res 2020;24:91–98.

12. Fagon JY, Chastre J, Wolff M, Gervais C, Parer-Aubas S, Stéphan F, Similowski T, Mercat A, Diehl JL, Sollet JP, Tenaillon A. Invasive and noninvasive strategies for management of suspected ventilator- associated pneumonia. A randomized trial. Ann Intern Med 2000;132.:

13. Fujitani S, Cohen-Melamed MH, Tuttle RP, Delgado E, Taira Y, Darby JM. Comparison of semiquantitative endotracheal aspirates to quantitative non-bronchoscopic bronchoalveolar lavage in diagnosing ventilator-associated pneumonia. Respir Care 2009;54.:

14. Ranzani OT, Senussi T, Idone F, Ceccato A, Bassi GL, Ferrer M, Torres A. Invasive and non-invasive diagnostic approaches for microbiological diagnosis of hospital-acquired pneumonia. Crit Care 2019;23:1–11.

15. el-Ebiary M, Torres A, González J, de la Bellacasa JP, García C, Jiménez de Anta MT, Ferrer M, Rodriguez-Roisin R. Quantitative cultures of endotracheal aspirates for the diagnosis of ventilator- associated pneumonia. Am Rev Respir Dis 1993;148:1552–1557.

16. Tran K, Cimon K, Severn M, Pessoa-Silva CL, Conly J. Aerosol Generating Procedures and Risk of Transmission of Acute Respiratory Infections to Healthcare Workers: A Systematic Review. PLoS One 2012;7.:

17. Wahidi MM, Lamb C, Murgu S, Musani A, Shojaee S, Sachdeva A, Maldonado F, Mahmood K, Kinsey M, Sethi S, Mahajan A, Majid A, Keyes C, Alraiyes AH, Sung A, Hsia D, Eapen G. American Association for Bronchology and Interventional Pulmonology (AABIP) Statement on the Use of Bronchoscopy and Respiratory Specimen Collection in Patients with Suspected or Confirmed COVID-19 Infection. J Bronchology Interv Pulmonol 2020;doi:10.1097/LBR.0000000000000681.

18. Wahidi MM, Shojaee S, Lamb CR, Ost D, Maldonado F, Eapen G, Caroff DA, Stevens MP, Ouellette DR, Lilly C, Gardner DD, Glisinski K, Pennington K, Alalawi R. The Use of Bronchoscopy During the Coronavirus Disease 2019 Pandemic: CHEST/AABIP Guideline and Expert Panel Report. Chest doi:10.1016/j.chest.2020.04.036.

19. Torrego A, Pajares V, Fernández-Arias C, Vera P, Mancebo J. Bronchoscopy in Patients with COVID-19 with Invasive Mechanical Ventilation: A Single-Center Experience. Am J Respir Crit Care Med 2020;202:284–287.

20. Kon ZN, Smith DE, Chang SH, Goldenberg RM, Angel LF, Carillo JA, Geraci TC, Cerfolio RJ, Montgomery RA, Moazami N, Galloway AC. Extracorporeal Membrane Oxygenation Support in Severe COVID-19. Ann Thorac Surg 2020;doi:10.1016/j.athoracsur.2020.07.002.

21. Singer BD, Jain M, Budinger GRS, Wunderink RG. A Call for Rational Intensive Care in the Era of COVID-19. Am J Respir Cell Mol Biol 2020;63.:

22. Kellerman SE, Herold J. Physician response to surveys. A review of the literature. Am J Prev Med 2001;20:61–67.

